# Favorable outcome on viral load and culture viability using Ivermectin in early treatment of non-hospitalized patients with mild COVID-19 – A double-blind, randomized placebo-controlled trial

**DOI:** 10.1101/2021.05.31.21258081

**Authors:** Asaf Biber, Michal Mandelboim, Geva Harmelin, Dana Lev, Li Ram, Amit Shaham, Ital Nemet, Limor Kliker, Oran Erster, Eli Schwartz

## Abstract

**Background:** Ivermectin, an anti-parasitic agent, also has anti-viral properties. Our aim was to assess whether ivermectin can shorten the viral shedding in patients at an early-stage of COVID-19 infection.

**Methods:** The double-blinded trial compared patients receiving ivermectin 0·2 mg/kg for 3 days vs. placebo in non-hospitalized COVID-19 patients. RT-PCR from a nasopharyngeal swab was obtained at recruitment and then every two days. Primary endpoint was reduction of viral-load on the 6^th^ day (third day after termination of treatment) as reflected by Ct level>30 (non-infectious level). The primary outcome was supported by determination of viral culture viability.

**Results:** Eighty-nine patients were eligible (47 in ivermectin and 42 in placebo arm). Their median age was 35 years. Females accounted for 21·6%, and 16·8% were asymptomatic at recruitment. Median time from symptom onset was 4 days. There were no statistical differences in these parameters between the two groups.

On day 6, 34 out of 47 (72%) patients in the ivermectin arm reached the endpoint, compared to 21/ 42 (50%) in the placebo arm (OR 2·62; 95% CI: 1·09-6·31). In a multivariable logistic-regression model, the odds of a negative test at day 6 was 2.62 time higher in the ivermectin group (95% CI: 1·06–6·45). Cultures at days 2 to 6 were positive in 3/23 (13·0%) of ivermectin samples vs. 14/29 (48·2%) in the placebo group (p=0·008).

**Conclusions:** There were significantly lower viral loads and viable cultures in the ivermectin group, which could lead to shortening isolation time in these patients.

**The study is registered at ClinicalTrials.gov NCT 044297411.**

## Background

Ivermectin is an FDA-approved broad spectrum anti-parasitic agent, which was initially approved in humans in 1987 to treat onchocerciasis, awarding the discoverers the Nobel prize of Medicine in 2015. Its main activity was known for therapy against infections caused by roundworm parasites. Over the years, the spectrum was extended and included also parasitic skin infections such as scabies among others. [1]

In the last decade, several in-vitro studies have shown its anti-viral activity against a broad range of viruses, mainly RNA viruses including HIV, influenza and several flaviviruses such as Dengue virus (DENV), Zika, and West Nile Virus.[2-6] Recently ivermectin was tested in vitro against SARS-CoV-2 and showed ∼5000-fold reduction (99.8%) in viral RNA after 48 hours.[7] However, it was criticized that the dosing used in the study cannot be achieved with the current approved dose.[8]

In addition, ivermectin has anti-inflammatory properties.[9] Since the excessive inflammatory response to SARS-CoV-2 is thought to be a major cause of disease severity and death in patients with COVID-19, [10] ivermectin may have further value in addition to its anti-viral properties.

With its high safety profile, ivermectin is a potential treatment against COVID-19 in its different stages. Some clinical studies have shown beneficial results regarding clinical outcomes and the length of viral shedding, however most of them are lacking a high standard of rigorous methodology.[11]

Here we conducted a double blinded randomized control trial to assess whether Ivermectin can shorten viral shedding, in non-hospitalized patients at the early stage of COVID-19 infection.

## Methods

### Study design

A randomized controlled, double blinded trial to evaluate the effectiveness of ivermectin in reduction of viral shedding among mild to moderate COVID-19 patients. The study was conducted in hotels located in (Tel-Aviv, Jerusalem and Ashkelon) Israel, that have been designated as isolation facilities for mild to moderate COVID-19 patients, not requiring oxygen.

Institutional Review Board (IRB) approval was given by the Sheba Medical Center’s IRB (7156/20). Written informed consent was received from each participating individual before recruitment.

### Study population

Patients were eligible for enrollment in the study if they were 18 years of age or older; not pregnant; with molecular confirmation of COVID-19 by RT-PCR; and with the intention to receive results within the first three days from symptoms onset. However due to the delay (three to four days) in getting results in the community, we extended the time up to seven days from symptoms onset. Since our main outcome was the change in viral shedding (as reflected by Ct value), asymptomatic cases were also included within 5 days from molecular diagnosis.

Patients were excluded if they weighed below 40kg, were with known allergy to the drugs, unable to take oral medication, participating in another RCT for treatment of COVID-19. In addition, patients who had RT-PCR results with Ct (cycle threshold) value >35 in first two consecutive were excluded. Patients with comorbidities of cardiovascular disease, diabetes, chronic respiratory disease (excluding mild intermittent asthma), hypertension, and or cancer were defined as high-risk patients.

### Randomization

Randomization in a 1:1 ratio was done by computer-generated program using randomization.com (http://www.jerrydallal.com/random/randomize.htm) by Clinical Research Coordinator (CRC), blinded to the rest of the study team. This CRC was not requiting patients and the numbered pills bottles were available only for the physician who were requiting.

Patients assigned to the intervention arm received ivermectin in a dosage regimen according to body weight; patients weighing between 40-69 kg received four tablets (=12mg) daily and patients weighing ≥70kg received five tablets (=15mg) daily, all for three days. Patients assigned to the placebo arm received the same number and same appearance of pills per weight daily, for three days. They were guided to take the pills one hour before a meal. The investigators and patients were blinded to the assignment.

### Intervention

On the day of randomization and treatment initiation, patients were tested for SARS-CoV-2 by reverse transcriptase polymerase chain reaction (RT-PCR) from nasopharyngeal (NP) swabs (day zero). Tests were then administered every two days from day six up to day 14, unless patients were discharged earlier from the isolation facilities. The protocol was amended at the beginning of September when the Ministry of Health changed the policy of isolation and allowed infected patients to leave the facility 10 days from symptom onset without further testing. At this point testing at day two and four were added to the protocol.

Sinc results of the test could have been influenced by the examiner who performed the swab and with differences between labs,[12, 13] a small number of trained practitioners were allocated to obtain the swab during the entire rial and instructed to use a uniform technique. In addition, all RT-PCR tests, including verification that patients were positive on day zero, were conducted by the same lab, at the Israel Central Virology Laboratory of the Ministry Of Health (located at Sheba Medical Center).

Patients were followed up daily by telephone until their discharge. Patients were asked whether they took the pills as guided, if they noticed any adverse effect following treatment and whether there were any follow up of symptoms.

Unexpectedly some patients who were isolated in the hotels as verified positive patients were found to be borderline or negative upon our RT-PCR test (Figure 1). Therefore,

**Figure 1:**
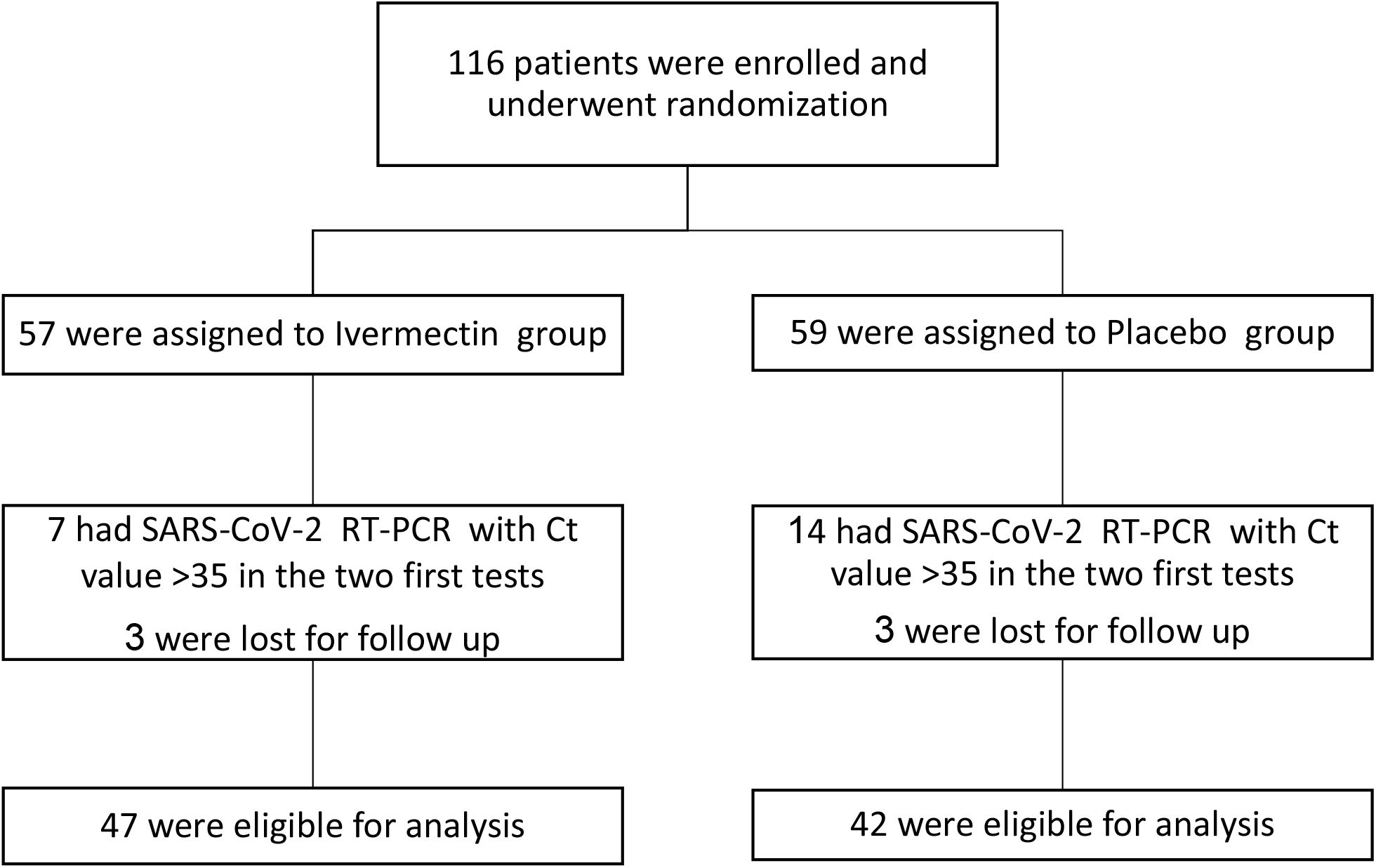
Enrollment and patient flow

Patients with missing data along the follow up were carried over from the last data available

### Outcomes

The primary clinical endpoint was viral clearance following a diagnostic swab taken on the sixth day (third day after termination of treatment), in the intervention group compared to placebo. Although negative PCR is defined in Israel with Ct>40, reaching this level may take a few weeks, and there is significant evidence that a non-infectious state is usually achieved at Ct level>30.[14-16] Therefore we defined a negative test at a non-infectious level as measured by RT-PCR of Ct values >30 (less than 3·4×10^4^ viral copies)

#### Post-Hoc analysis

Toward the end of our study (January 2021) the central virology lab. established a Biosafety Level 3 (BSL-3) unit, allowing us the ability to culture the virus. Since positive medium of participating patients were kept in -80c, we were enabled to culture them. Thus, an end point of culture viability at days 2-6 post-intervention was added.

### PCR testing

The presence of the SARS-COV-2 RNA was detected using the Seegene Allplex CoV19 detection kit, according to the manufacturer’s instructions (See supplement). The test detects three viral genes: envelop (E), nucleocapsid (N) and RNA-dependent RNA polymerase (RdRp). For each sample, Ct level was defined as the Ct level of the highest viral load (low Ct).

### Viral copies Determination

Determination of the copies number in the examined samples was performed by generating standard curves for each reaction, thereby enabling the conversion of the Cq values to viral genome copies, as detailed in the Supplementary methods.

### In-Vitro cultures

Positive samples were stored at -80°C and were thawed for culturing on Vero E6 cells at 37°C for seven days, as detailed in the Supplementary methods.

### Statistical methods

#### Sample size

Based on published data from the Ministry of Health at the time of study initiation, we expected less than 10% of patients at day six show a negative RT-PCR test. With the interventional drug we expected a reduction of at least 25% in the proportion of positive cases. Hence, considering a potential decrease from 90% to 67·5% (25% decrease), with a power (1-β) of 80% at a significance level of 5% (α= 0·05), a minimal sample size of 96 participants in total, was required to detect a statistically significant difference. Therefore, 48 patients were needed in each study arm. To account for a loss to follow-up of 10% after 14 day, we aimed to recruit a total of 105 participants.

#### Statistical analysis

Statistical analysi**s** was done by the Biostatistics and Biomathematics Unit, Gertner Institute, Sheba Medical Center, Tel-Hashomer, Israel.

Continuous variables are presented as mean ± standard deviation or as median and interquartile range. Categorical variables are presented as N (%). Differences between ivermectin and placebo groups were assessed using a Chi-square test and t-test, for categorical and continuous data respectively. Where cross tabulation frequencies were less than 5, the Fisher exact test was used. A multivariate logistic regression model was used to determine the impact of ivermectin while controlling for age, sex, weight, and being symptomatic or not on reduction of viral load on day 6^th^ as reflected by Ct level>30. Results include adjusted odds ratios (OR), and 95% confidence intervals (CI). Kaplan-Meier curves were drawn, and survival analysis conducted with log-rank test using for time to negative RT-PCR (Ct level>30) result.

Boxplots were produced in R version 4·0·2. For figure readability, viral load values were log-transformed.

For all analyses, significance was set at p < 0·05. All data analyses were performed with the SAS 9·4 software (Cary, NC, USA).

## Results

From May 15^th^, 2020 through end of January 2021, a total of 116 patients underwent randomization, and 89 were eligible for analysis (Figure 1).

The baseline study population characteristics are detailed in Table 1. The median age of the patients was 35 years (range, 20 to 71), 22·4% equal or older than 50 years and 7·8% equal or older than 60 years. A majority of the patients were males (78·4%). Twelve (13·5%) patients had comorbidities associated with risk for severe disease [17]; 17% and 9·5% among the ivermectin and placebo groups respectively, p=0·53.

**Table 1:** Baseline study population.

|  | **All (N=89)** |  | **Ivermectin group**<br>**(N=47)** |  | **Placebo group**<br>**(N=42)** |  | **P\*** |
| --- | --- | --- | --- | --- | --- | --- | --- |
| Male gender *n*, (%) | 69 | (78.4) | 36 | (78.3) | 33 | (78.6) | 0.9718 |
| Age *median (IQ range)* | 35.0 | (28.0-47.0) | 36.0 | (32.0-50.0) | 33.5 | (26.0-47.0) | 0.2446 |
| Weight *median (IQ range)* | 79.0 | 70.0-86.0) | 80.0 | (70.0-90.0) | 75.0 | (67.0-85.0) | 0.2545 |
| Symptomatic *n*, (%) | 72 | (80.9) | 37 | (78.7) | 35 | (83.3) | 0.5807 |
| Days from symptoms onset** *median (IQ range)* | 4.0 | (3.0-5.0) | 4.0 | (3.0-5.0) | 4.0 | (3.0-5.0) | 0.9170 |
| Ct value on day 0 *median (IQ range)* | 23.0 | (20.0-28.0) | 24.0 | (21.0-28.0) | 22.0 | (19.0-27.0) | 0.1030 |

\*P value by Fisher exact test for categorical variables or by Kruskal–Wallis test for continuous variables
\*\* calculated for only for symptomatic patients

### Clinical Presentation

Among the study population 83% were symptomatic. The most common symptoms, fatigue, fever, cough, headache, and myalgia were prevalent in approximately half of the study population. (symptoms detailed in Table S1-supplement). None of these variables were statistically different between the two study arms.

### Study Outcome

Viral load during the study period is depicted in Figure 2. Viral load of the ivermectin group decreased faster in comparison to the placebo group at the early stage of the intervention, during days two to six. As spontaneous recovery takes place also in the placebo group the viral load is decreasing, having similar viral load since day eight.

**Figure 2:**
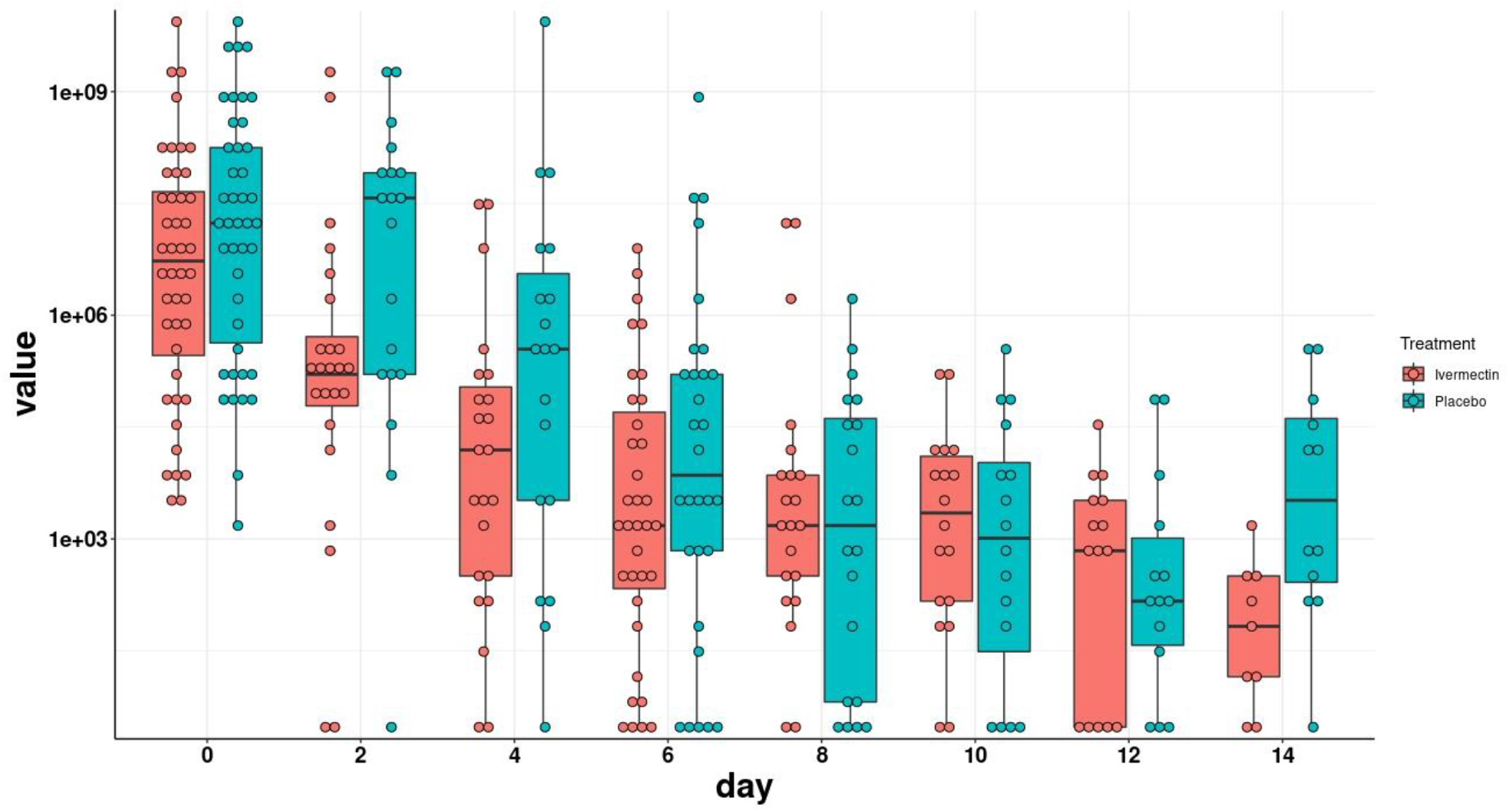
Viral load evolution by study arm. Viral load values were log-transformed. The boxes show the interquartile range. Dots represent each individual value.

As mentioned above, our calculations were based on negative results reflected in Ct >30. The rate of negative RT-PCR for SARS-CoV-2 at day four (one day after termination of treatment) through day ten was higher in patients receiving ivermectin but was statistically significant on days six to eight (Table 2).

**Table 2:** The negative RT-PCR (Ct>30) test for SARS-CoV-2 results, ratio at days 4 to 10.

| A. Based on RT-PCR (Ct>30) test |  |  |  |  |  |  |  |
| --- | --- | --- | --- | --- | --- | --- | --- |
|  | N | Ivermectin | Placebo | P value* | OR | 95% CI |  |
| Day 4 | 50 | 15/28 (54%) | 7/22 (32%) | 0.12 | 2.47 | 0.77 | 7.92 |
| Day 6 | 89 | 34/47 (72%) | 21/42 (50%) | 0.03 | 2.62 | 1.09 | 6.31 |
| Day 8 | 89 | 39/47 (83%) | 25/42 (59%) | 0.01 | 3.32 | 1.25 | 8.82 |
| Day 10 | 89 | 40/47 (85%) | 29/42 (69%) | 0.07 | 2.56 | 0.91 | 7.72 |
| B. Based on RT-PCR (Ct>30) test together with non-viable cultures |  |  |  |  |  |  |  |
| Day 4 | 50 | 24/28 (86%) | 13/22 (59%) | 0.03 | 4.1538 | 1.0688 | 16.1439 |
| Day 6 | 89 | 44/47 (94%) | 31/42 (74%) | 0.01 | 5.2043 | 1.3400 | 20.2129 |
| Day 8 | 89 | 45/47 (96%) | 32/42 (76%) | 0.01 | 7.0313 | 1.4419 | 34.2875 |
| Day 10 | 89 | 45/47 (96%) | 36/42 (86%) | 0.14** | 3.7500 | 0.7135 | 19.7078 |

\*P value by Chi square test , \*\*P value by Fisher exact test

In the multivariable logistic regression model, the adjusted odds ratio of negative SARS-CoV-2 RT-PCR nega
tive test (Ct>30) at day six and eight for the ivermectin group were 2·62 (95% CI: 1·06–6·45, P=0·04) and 3·87 group (95% CI: 1·36–11·04, P=0·01) fold higher than for the placebo group, respectively. (Table 3)

**Table 3:**
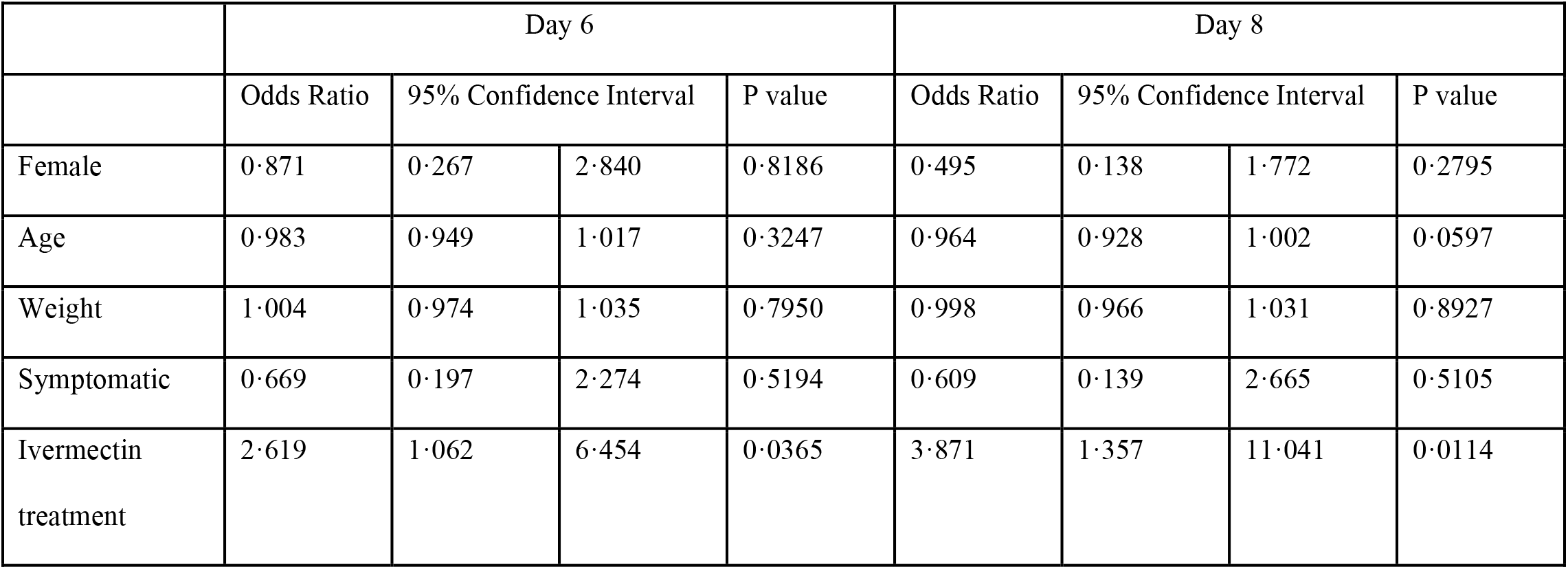
Multivariable analysis for negative RT-PCR (Ct>30) test for SARS-CoV-2 results on day 6 and 8.

Kaplan-Meier analysis (Figure 3) adjusted to symptom onset showed the significant difference between the ivermectin and placebo arms during the course of treatment.

**Figure 3:**
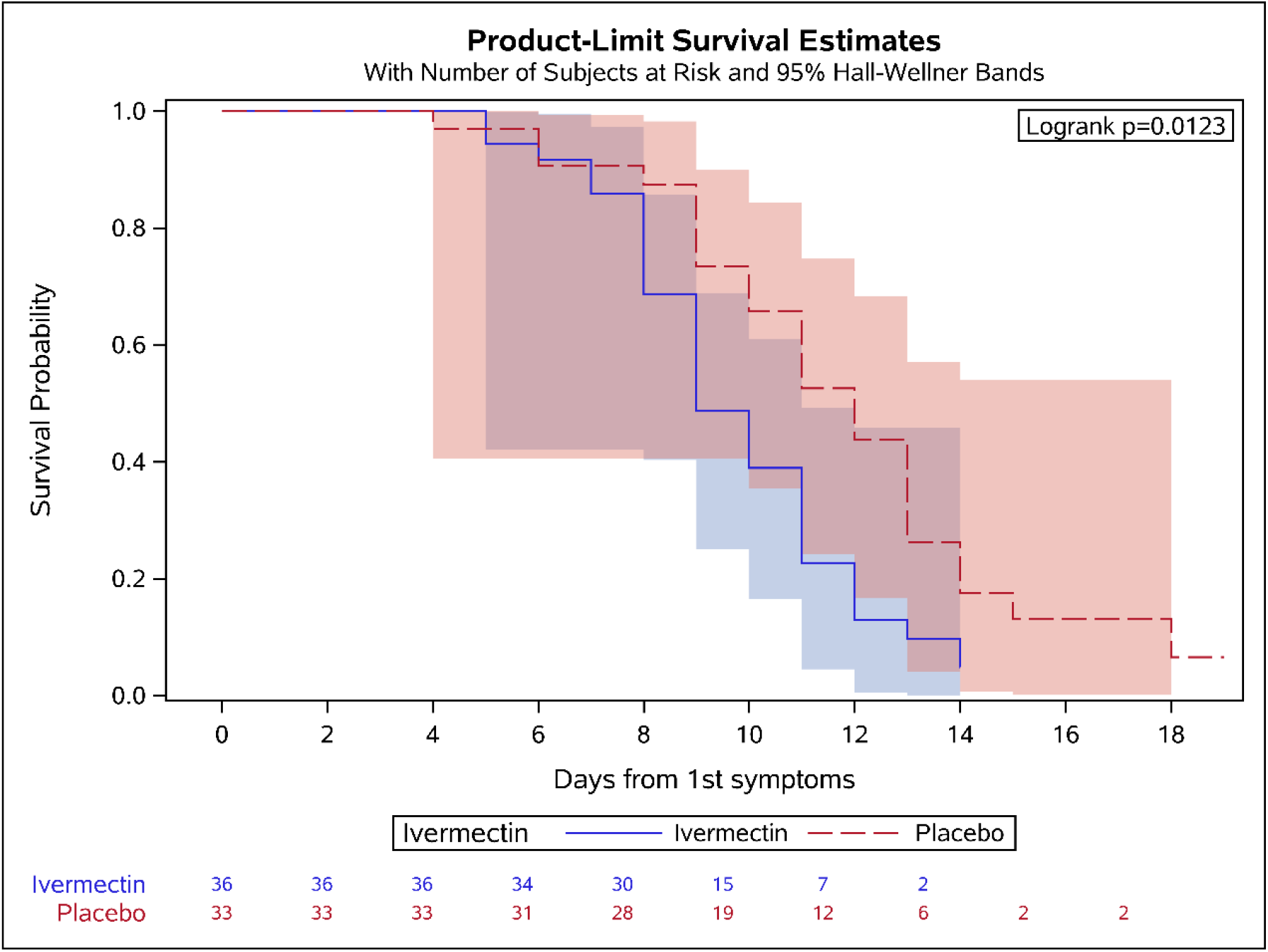
Kaplan-Meyer curve for time to negative (Ct <30) results for symptoms onset (calculation is done for symptomatic patients, N=69)

### Clinical outcome

During the study period four patients were referred to hospitals, with three of them being in the placebo arm. The first placebo-treated patient was hospitalized for 11 days with prolonged respiratory symptoms and needed oxygen even after his discharge from hospital. The second was hospitalized for one day due to respiratory complaints. The third one was referred to hospital due to headache and dizziness and was diagnosed with sinusitis after evaluation (brain CT and MRI). In addition, one asymptomatic patient became symptomatic, which occurred in the placebo group. In the ivermectin arm, one patient was referred to hospital due to shortness of breath at the day of recruitment. He continued the ivermectin and a day later was sent back to the hotel in good condition.

### Culture Positivity rate

A convenient sample of 16 samples on day of recruitment (day zero) was cultured. Ct levels ranged from 14-28 (mean 21·5 ± 4·1), and among them 13/16 (81·2%) turned to be positive. Culture viability was tested further by available samples on days two, four and six after intervention (see details Table S2–supplement). Altogether 52 samples were cultured; viable culture in the placebo group were positive in 14 out of 29 cultures (48·2%) while among the ivermectin group, only 3/23 (13·0%) were found positive (P=0·008).

In a composite calculation, taking into account Ct values >30 together with non-viable culture, the negative results of the ivermectin group reached significance even at day four (one day after ending the treatment) with 86% negative patients compared to 59% in the placebo group (P=0·04) (see Table 2b).

### Adverse events

Among the 116 intention to treat participants, 3 patients reported having diarrhea following the treatment, two (3·5%) in the ivermectin group and one (1·7%) in the placebo group. In all cases the diarrhea resolved in two days. Two patients in the placebo arm reported rash during the treatment course which subsided within one to two days. No other adverse effects were reported. All of the eligible 89 patients for analysis reported to be adherent to the treatment as guided.

## Discussion

In this double-blind, randomized trial with mild COVID-19 patients, ivermectin significantly reduced time of viral shedding and affected viral viability when initiated at the first week after evidence of infection. Our primary endpoint was to show the benefit of ivermectin on day six (three days after ending treatment) which was achieved with 72% of samples being non-infectious (Ct>30) in comparison to 50% among the placebo group (OR 2·6). Even at day 4 (1 day after treatment end) the ivermectin group showed an OR of 2·4, although this did not reach significance, possibly due to a small sample size on that day.

The anti-viral activity was also reflected in the Kaplan-Meier curve where the effect of the drug was seen after the second day of treatment (Figure 3).

To further explore the anti-viral activity, we observed the culture viability in both placebo and ivermectin groups. This post-hoc analysis became available in our institution at the end of the study only, when the BSL-3 lab was established (January 2021). The results show the advantage of ivermectin where only 13% of samples stayed positive on days 2two to six, while 48% stayed positive in the placebo group (P=0·008).

The broad-spectrum antiviral activity of ivermectin is related to its ability to target the host importin (IMP) α/β1 nuclear transport proteins responsible for nuclear entry of cargoes of viral proteins, which in turns block the host anti-viral activity. In some viruses the viral protein (such as integrase and NS5) has been identified while in SARS-CoV-2 the protein was not identified.[18, 19] The anti-viral properties of ivermectin against SARS-CoV-2 was shown in an in-vitro model.[7] A major criticism regarding this in-vitro model was that the ivermectin concentration used was more than 35 times higher than the maximum plasma concentration after oral administration of the approved dose.[8] But higher doses may not be necessary as some models predict that the lungs achieve higher concentrations, up to 10-fold higher than in the serum.[20] In addition, ivermectin concentrations in blood may not reflect the activity of its other metabolites which might be the active agents.[21, 22]

In humans, several randomized control trials have been recently published. A double-blind randomized control trial conducted in Colombia by López-Medina et al., included 476 mild patients randomly assigned to receive either oral ivermectin 300 mcg/kg for five days vs. placebo.[23] The study was initially aimed to examine ivermectin as an agent which could prevent clinical deterioration when given during the early stage of COVID-19. However, the endpoint of this study was altered to be time to resolution of symptoms within 3 weeks, as the original endpoint of clinical deterioration could not be achieved due to the low number of hospitalizations in their cohort. As the authors mentioned in their limitations, reduction in the viral load or viral shedding better reflects the anti-viral activity of the drug rather than longevity of symptoms[23]. Peer reviewed randomized control trials from Bangladesh support our finding of faster viral clearance in the ivermectin group, and in a small RCT from Spain, treatment with ivermectin showed a tendency toward faster viral clearance.[24, 25]

One may wonder about the public health benefit of treating mild patients since shortening the period of relatively mild symptoms may not merit a mass drug administration of ivermectin to the large population of non-hospitalized patients. However, inducing faster viral clearance and therefore reducing the time until the patient reaches a state of being non-infectious have an extremely important public health impact. Taking the two composites; Ct values above 30 and negative cultures, leads to almost 90% non-infectious status at day four (one day after ending treatment) among ivermectin users []. From the public health point of view, it may shorten isolation time, which can serve as a major relief of the economic and social burden.

Our study has several limitations. First, the sample size was relatively small, and was designed to look for differences in viral load, but not for clinical deterioration and prevention of hospitalization. The second limitation was that drug therapy was not physically observed by investigators. Finally, our study was conducted among mild-non-hospitalized patients and therefore the results cannot be applied to a more severe or immune-suppressed populations.

The strength of our study was its double-blind structure with more substantial outcomes such as Ct values and culture viability where the laboratory personnel did not have any information concerning the patients’ assignment. In conclusion, our study strongly supports the notion that ivermectin has anti-SARS-CoV-2 activity. If used at the early stage of disease onset, it may shorten the isolation time and reduce transmission. Further studies are needed to test its ability to prevent clinical deterioration for high-risk groups and to examine its potential as a prophylactic drug. Vaccines are now starting to become available, but it will take years before they are distributed worldwide. As this drug may also reduce mortality, urgent intervention with further well-designed studies are needed.

## Data Availability

All data supporting the results will be provided by the corresponding author upon publication.

## Acknowledgement

We would like to thank Super-Pharm Professional for donating the drug and the placebo pills, Ms. Liraz Olmer for statistical analysis support, Ms. Rivka Goldis for administration aids, Dr. Emiliano Cohen for graph producing, and Mr. Nadav Cain for his logistic support. Finally we would like to thank the Dirctorate of Defense Research and Development (DDR&D) at Israel’s Ministry of Defense and Home Front Command staff who helped us accessing the dedicated corona hotels, without their support the study could not been performed.

## Access to data

All data supporting the results will be provided by the corresponding author upon publication.

## Contibutions

Eli Schwartz and Asaf Biber had full access to all of the data in the study and take responsibility for the integrity of the data and the accuracy of the data analysis.

Conceptualization: ES

Data curation: ES, AB, MM

Formal analysis: ES, AB, MM, OE

Investigation: AB, MM, GH, DL, LR, AS, IN, LK, OE, ES

Methodology: ES, AB, MM, OE

Supervision: ES, AB

Writing - original draft: ES, AB, DL

Writing - review & editing: all authors contributed, reviewed and approved the last draft.

