## Supplement for "Favorable outcome on viral load and culture viability using Ivermectin in early treatment of non-hospitalized patients with mild COVID-19 – A double-blind, randomized placebo-controlled trial"

### PCR testing

NP swabs were inactivated by 10 minutes incubation with Lysis buffer (PrimeStore® Molecular Transport Medium or Zymo DNA/RNA Shield, https://www.zymoresearch.com/). Total RNA was extracted using the MagNA Pure 96 system (ROCHE, https://lifescience.roche.com) or the PSS MagLEAD extractor (http://www.pss.co.jp), according to the manufacturer's instructions. The presence of the SARS-COV-2 RNA was detected using the Seegene Allplex CoV19 detection kit, according to the manufacturer's instructions (http://www.seegene.com/assays/allplex_2019_ncov_assay). Briefly, the test detects three viral genes: envelop (E), nucleocapsid (N) and RNA-dependent RNA polymerase (RdRp). The integrity of the extraction procedure is monitored using an internal control that is inserted into the sample prior to the extraction procedure. The PCR integrity is monitored using a CoV19 positive control. Following mix assembly, the samples were analyzed using the Bio-Rad CFX96 thermal cycler, and its accompanying software, CFX Maestro (https://www.bio-rad.com/).

For each sample, Ct level was defined as the Ct level of the highest viral load (low Ct).

**Viral Copies:** Determination of the viral genome copy number in each sample was done by generating standard curves for each reaction, which allows for conversion of the PCR Cq values to target copies. PCR-generated targets for each reaction in the Allpex test were transcribed to RNA using the MegaScript T7 kit (www.thermofisher.com/). The resulting RNA of each target was purified and its concentration was measured for each target. The number of target copies in 1 ng of purified RNA was calculated based on the amplified sequence length, thereby enabling the conversion from copy number to Ct values. Serial dilutions of each target amplicon were used for RT-qPCR to generate the standard curve for each reaction. The Ct values were then plotted against the calculated concentration, using a semi-logarithmic scale graph, and the derived regression formula was calculated. The Ct values for each sample were converted to viral genome copies using the resulting formula.

**I-Vitro cultures:**

Vero E6 cells were cultured at 37°C until reaching 70% confluence. Infection was performed using samples that tested positive SARS-CoV-2 in the PCR assay with Ct value <30. The culture was incubated for 1 hour at 33ºC with 300 µl of the filtered nasopharyngeal samples (filtration is performed to prevent bacterial infection of the culture), followed by addition of 5 ml 2% FCS MEM-EAGLE medium. The infected cells were cultured for 7 days and were inspected visually for the onset of cytopathic effect (CPE). Upon CPE onset, supernatants were collected, and RNA was extracted as described above. The viral propagation load was determined by SARS-COV-2-specific RT-qPCR, as described above.

**Table S1:** **<u>Symptoms at presentation</u>**

| Symptom | All | Ivermectin | Placebo |
|---|---|---|---|
| Fatigue | 56·8 | 55·3 | 57·1 |
| Headache | 50·6 | 48·9 | 52·4 |
| Cough | 49·4 | 48·9 | 50 |
| Fever | 48·3 | 51·1 | 45·2 |
| Myalgia | 42·7 | 42·6 | 42·9 |
| Sore throat | 30·3 | 27·7 | 33·3 |
| Lss of taste | 28·1 | 34 | 21·4 |
| Loss of smell | 28·1 | 34 | 21·4 |
| Dizziness | 19·1 | 21·3 | 16·7 |
| Chest pain | 19·1 | 17 | 21·4 |
| Asymptomatic | 16·8 | 20 | 14·3 |
| Dyspnea | 15·7 | 19·1 | 11·9 |
| Nausea | 15·7 | 17 | 14·3 |
| Abdominal pain | 14·6 | 21·3 | 7·1 |
| Diarrhea | 12·3 | 10·6 | 14·3 |
| Vomiting | 4·5 | 2·1 | 7·1 |
| **Rhinorrhea** | **2·2** | 2·1 | 2·4 |
| Backpain | 2·2 | 2·1 | 2·4 |
| Rash | 1·1 | 0 | 2·4 |
| Horseness | 1·1 | 0 | 2·4 |

**Table S2: Culture viability at 2-4- 6 days after initiating treatment**

| | Day 2 Positive/Tested | Day 4 Positive/Tested | Day 6 Positive/Tested | Total Positive/Tested* (%) |
|---|---|---|---|---|
| Ivermectin | 2/12 | 1/5 | 0/6 | 3/23 (13·0%) |
| lacebo | 8/14 | 5/10 | 1/5 | 14/29 (48·2%) |

*Fisher-exact test - P value = 0·008
